# Enhancing Food Security for Families Vulnerable to COVID-19

**DOI:** 10.1101/2021.02.04.21251181

**Authors:** Tayo Fabusuyi, Aditi Misra, Jonatan Martinez, Andong Chen, Vivian Jiang, H. V. Jagadish, Robert C. Hampshire

## Abstract

The COVID-19 pandemic has exposed some of the underlying inequities in our society. Vulnerable and low-income communities who have typically struggled with food insecurity have been further impacted by the COVID-19 pandemic. To make matters worse, transit dependent households in the City of Detroit that rely on SNAP benefits from the government have a 54% probability of not having internet access, reducing the possibility of online food delivery. Thus, the motivation for the Detroit Food Delivery Pilot Program, a prepared meal delivery service for food insecure households during the COVID-19 pandemic. Through this program, the City had aided 350 households; over 1,000 unique individuals and has served over 90,000 meals. This had all been made possible through the engagement of 575 volunteers, volunteering over 6,850 delivery miles. Our team helped to identify the magnitude and the needs of the City’s food insecure population; then partnered with the City of Detroit to modify existing food programs to address the crisis, and finally, assessed the impact of the program on the target population. Our analysis revealed that approximately 20,800 households will be classified as food insecure in Detroit and only a fraction of this population is benefitting from the food delivery pilot program. However, on average, the concerns about food sufficiency have abated appreciably from the surveys analyzed. Responses from the beneficiaries of the food delivery pilot program were positive though concerns about the travel behavior of individuals, particularly those from COVID-19 positive households remain.

## 1 INTRODUCTION AND BACKGROUND CONTEXT

The COVID-19 pandemic has exposed some of the underlying inequities in our society. Excluding health outcomes, nowhere is this impact more pronounced and consequential than access to food. Vulnerable and low-income communities who have typically struggled with food insecurity have been further impacted by the COVID-19 pandemic. The closure of food service outlets or the movement to an “order delivery” business mode during the shut-down period in many states, coupled with reduced public transit services have severely affected transit dependent households. The situation was made more dire by the fact that within the City of Detroit, transit dependent households that rely on the Supplemental Nutrition Assistance Program (SNAP) from the government have a 54% probability of not having access to the internet. This precludes them from using the services of food delivery services such as Postmates, Grubhub, Doordash or UberEats. In addition, access to school lunch programs that children in low income families often depend on, may no longer be available given widespread school closures and the movement of classroom instructions to the virtual space.

The above issues explain the primary motivation for this project: the Detroit Food Delivery Pilot Program, a prepared meal delivery service for food insecure households. Beginning at the height of the COVID-19 crisis, and with funding from the National Science Foundation (NSF), we partnered with the City of Detroit to identify the needs of the City’s food insecure population. We provided input on identifying the target population and estimating the magnitude of food insecure households within the city, the modification of existing programs to address the crisis situation, as well as an assessment of the degree to which the program as implemented positively impacted the target population. These tasks were carried out using a myriad of datasets including secondary data from the USDA on food insecurity, social vulnerability index from the Centers for Disease Control (CDC), the US Census American Community Survey (ACS) and Public Use Microdata Sample (PUMS) dataset; document review from the City of Detroit, particularly Detroit Department of Transportation (DDOT) and primary data from surveys administered on food delivery programs.

Given the need to pick up the meals from where they are prepared, and then be then delivered to households in need, the project falls within the broader umbrella of urban freight transport [1], [2], [3]. A decent body of work exists in this area especially regarding “Meals on Wheels” food delivery programs that typically target seniors and individuals with disabilities [4], [5], [6] with some of the literature focusing on the routing of the meals primarily from an operations research perspective [7], [8], [9]. In contrast to a primarily elderly Meals on Wheels client base, the target population of this project typically consists of multi-generational, low income and transit dependent households. Because of the pandemic, access to food by this segment of the population has been further restricted given reduction in hours of grocery stores, more limited public transportation as well as the increase in the volume of online food deliveries, driven largely by the closure of in-room dining by most restaurants during the early periods of the pandemic.

Individuals in these households may come from very different sociodemographic groups - from food insecure pupils dependent on school supplied lunches to seniors who do not have the tech savviness to order delivery via smartphone apps or to use online payment systems. The present research takes these issues into consideration by identifying food vulnerable populations and geographic areas affected using USDA data coupled with key sociodemographic variables while also reflecting COVID-19 case intensity data. The research effort seeks to document not only the outcome measures but also the process employed in identifying the population and in running the food delivery program.

The balance of the paper is organized as follows. Section 2 provides an historical context of food delivery programs focusing on those delivered through a partnership between the government and the nonprofit sector. The section also details the coordination mechanism employed in delivering the services. The Detroit Food Delivery Pilot Program is the focus of Section 3. Divided into three parts, Section 3 describes Detroit’s ecosystem of food delivery programs and documents both the design and implementation of the pilot program that is a response to the COVID-19 pandemic. Section 4 details the approach used for the analysis while the results are discussed in Section 5. Section 6, the last section, concludes and provides insights on potential extensions to the program.

## 2 SURVEY OF COMPARABLE FOOD DELIVERY PROGRAMS

A number of food delivery programs were created or adapted nationwide in response to COVID-19, with origins in a range of government, private sector, and non-profit organizations; some also arose from public-private partnerships. Prior to the onset of the pandemic, the majority of subsidized meal delivery services have catered to the elderly and/or disabled, or provided medically tailored or tailored meals to recipients of Medicaid and Medicare, depending on their plans. Of these services, the most prominent is Meals on Wheels (MOW), which supports the nutritional and social needs of homebound senior adults through meal delivery and in-home visits. MOW is an example of a public-private partnership: according to their 2019 fact sheet, the organization follows a hybrid-funding model, with the Older Americans Act (OAA) as a main source of funding from which 39% of their spending is sourced [10]. The rest of their funds are derived from a mix of “state and/or local sources, private donations from foundations, corporations and individuals, and federal block grants.”

In a study of the interactions between MOW drivers and clients at six different locations across the United States, it was observed that the majority of these programs relied very lightly on paid drivers: most of their deliveries were staffed by volunteers [11]. Recipients of meals have reported high levels of satisfaction with the service, emphasizing the additional benefits of improved health and increased independence in their own home. MOW staff and volunteers who deliver the meals also detail positives of the service, such the growth of strong social bonds and the perceived benefit of assisting clients with additional services, such as helping with small tasks around the house. Such externalities as these may be difficult to achieve in the current pandemic environment, as guidelines on social distancing and self-isolating heavily curtail person-to-person interactions.

MOW is an example of the broader service of Home Delivered Meals (HDM). Coordination of HDM services relies on numerous fluctuating factors such as volunteer supply and demand and available financial support for purchasing meal ingredients [9]. A case study of HDM planning conducted in Allegheny County, PA noted that federal subsidies for delivery operations have the provision that delivered meals must be at least 140°F; in an insulated carrier, this limits the delivery of a driver’s last meal to 45 minutes^13^. Restrictions like these factor into the location-routing problem (LRP) for home-delivered meals: time must be taken into account as kitchens in different locations decide how to transport meals to spatially dispersed customers. Routes are driven by multiple vehicles, and the number of routes assigned to each kitchen should not exceed the maximum allotted number. Meal recipients must be served on a single route by a single kitchen; additionally, kitchens cannot serve more than their maximum capacity of meals.

Operations research models for community-based programs like HDM also cover LRP issues for places like food banks: food bank vehicles work within the constraints of perishable food rescue programs and “the stop-and-drop” problem, where customers pick up food that is distributed to multiple operation sites [12]. The consideration of such procedures and route operations is pertinent to the current COVID-19 environment in which many new and existing community-based and government-backed programs must adapt to unique and uncertain circumstances.

New York City is currently providing access to free meals for all citizens. However, emergency home delivery is only available to individuals and families who meet a set of four criteria: no members can get food because they are at increased medical risk or are homebound; no neighbors or family members can help get food; there is no ongoing meal assistance from other providers; and there is an inability to afford meal or grocery delivery [12]. The deliveries may consist of pantry items, shelf stable meals, or fresh meals. Halal or Kosher food may also be requested. Each delivery contains 9 meals (three days of food); deliveries may also be recurring (nine meals twice a week) for up to three weeks.

Numerous organizations in the non-profit sector adapted their service models to better respond to the pandemic. For instance, Open Hand, a 501(c)(3) nonprofit in Atlanta, Georgia, provides a “Food is Medicine” program which delivers freshly prepared meals to at-risk youth, and medically-tailored meals to senior citizens in congregate settings and those transitioning from hospital care back home, including Medicaid clients. The onset of coronavirus led to Open Hand expanding their services statewide for the first time, first by shipping frozen meals to several rural counties across the state, then spreading to include middle and north Georgia, providing meals for “many of whom struggled with food insecurity even before the COVID-19 crisis.” An update from June 3rd on their website indicated an 26% increase in demand for their home-delivered meal service over the past two months, amounting to “an additional $75,000 meals” provided over that period [13].

In an example of public-private partnership, the city of Baltimore, Maryland partnered with Amazon to deliver grocery and produce boxes for the months of July and August. Those who qualify include households who “have no food at home or will run out before [they] can buy some more,” who cannot afford to pay for grocery or meal deliveries, or who are experiencing “financial, social, or medical hardships due to COVID-19” [14]. The partnership provides 30lbs of non-perishable food and 24lbs of fresh produce each week in July, as supplies last. Households that require food in August must call back at the beginning of that month to continue.

## 3 DETROIT FOOD DELIVERY PILOT PROGRAM

At inception, the Detroit Food Delivery Pilot Program, provided 2-3 weeks of contactless food delivery to households in the City of Detroit that had at least one individual who tested COVID-19 positive. These households consisted of both, residents who have previously faced food insecurity and live in communities challenged by inadequate transportation access, and those who, due to the impacts of pandemic (e.g. loss of employment and inability to be out in public), experienced novel cases of food and transportation insecurity. As the first wave of COVID-19 in the City of Detroit declined and COVID-19 positive households dropped below the capacity of the program, agencies who serve seniors, children, and other vulnerable populations were encouraged to refer families living at the intersection of food and transportation insecurity to the program. As COVID-19 positive cases increase and decrease with subsequent waves, it is important that the program adjust the population served to ensure all COVID-19 positive households can be served prior to other vulnerable households.

The program is, in part, a volunteer-based food delivery program. Volunteers are responsible for going to food distribution sites, where City Parks & Recreation staff load their trunks with food boxes that are to be distributed amongst two families. The volunteers then contact residents to inform them that they are on their way to drop off food boxes on their porches. Residents are instructed to stay indoors, to ensure contactless delivery. This program relies on its own pool of volunteers, who come out once a week to serve the Detroit community.

This program has relied on a patchwork of funding which has been changing throughout the duration of the program and as it shifts between different iterations. Initially, Parks & Recreation gymnasiums were repurposed to store, sort, and package the food for families. Staff, who would be otherwise furloughed, were assigned to receive, sort, and distribute food to drivers, and volunteers donated their time and vehicles to deliver the food from the distribution sites to the households in need. Food preparation is done primarily by Gleaners, Salvation Army, Isaiah Project, and Parks & Recreation while Parks and Recreation volunteers serve as the program’s public-facing distributors. All the food program activities are coordinated by a team of personnel from the City of Detroit’s Department of Innovation and Technology, Department of Parks and Recreation, and the University of Michigan Gerald R. Ford School of Public Policy.

The shelf-stable food was and will continue to be donated by the Salvation Army. The fresh produce boxes are and will continue to be, in part donated and in part bought from Gleaners, depending on need. Additionally, for approximately two months, the Isaiah Project provided pre-packed frozen meals for families with children. During the second iteration of the program, Detroit was able to allocate funds from the CARES Act and United Way to extend the duration of the program until the end of December 2020, expand the hours of operation, pay program staff, and to address other needs.

There are however issues that pose operational bottlenecks. There are two referral pipelines, the health department referral of COVID-19 positive households and the referral of vulnerable households from other stakeholders. Each stakeholder has its own criteria for determining which vulnerable households to referee to the program. However, at peak capacity, the program can serve 120 households a week, with priority given to COVID-19 positive households. This creates a situation where families who are in need but not COVID-19 positive, may not be served during waves of COVID-19 if the program’s capacity is not increased. In addition, the program relies on volunteers to deliver the food. The challenge is that volunteer participation rates fluctuate from week to week, creating scenarios where staff or others have to be tapped to volunteer. Having a more reliable source of transportation for weeks with low volunteer turnout can help address one of the major operational issues.

## 4 DATA SOURCES AND ANALYSIS

To understand the impact of COVID-19 on food insecure households in Detroit, we begin by posing the question “who are the households most vulnerable to the COVID-19 pandemic particularly with regards to food and in what areas of the City are they resident? We used data from the US Census; US Department of Agriculture (USDA) and by reviewing documents from Detroit department of Transportation (DDOT). In addition, we used secondary data obtained from the DMACS survey [15] to capture the severity and impact of the pandemic on this segment of the population with emphasis on the first wave of the survey which coincides with the shut-down period. Identifying the COVID-19 vulnerable population and assessing the severity of the disease on this population provided crucial information on modifying existing food programs to better respond to the needs. Finally, using primary data from surveys administered to patrons of the Detroit Food Delivery Pilot Program, we provide preliminary findings on the program’s effectiveness.

### 4.1 Identification of the population of food insecure households

In identifying the population vulnerable to COVID-19, we look at both the CDC’s social vulnerability index (SVI) and the USDA index on food security. The CDC defines socially vulnerable population as those “that are particularly vulnerable to disruption and health problems as a result of natural disasters, human-made disasters, climate change, and extreme weather”. The index ranges from 0 (least vulnerable) to 1 (most vulnerable) and it is made up of four categories of vulnerability – socioeconomic status, household composition and disability, minority status and language spoken at home, and housing and transportation. From merely eyeballing the map it is obvious that an appreciable part of the city is considered vulnerable. To enrich the thematic map, we have overlaid it with the locations of grocery stores across the city.

With regards the population of food insecure households, we examine the USDA data that is related to food access and that provides information on areas that could be regarded as food deserts within the city. Figure 1b provides a panoramic picture of access to food in Detroit in 2015, the most recent year for which data is available. Based on the USDA definition, a census tract (CT) is classified as a food desert if at least 500 residents, or 33% of its population, live more than a mile from a supermarket, supercenter or large grocery store. These CTs are visible throughout the city without being clustered in a location. Beyond the “500 or 33%” threshold, the severity of the lack of access is not captured by the measure.

**Figure 1:**
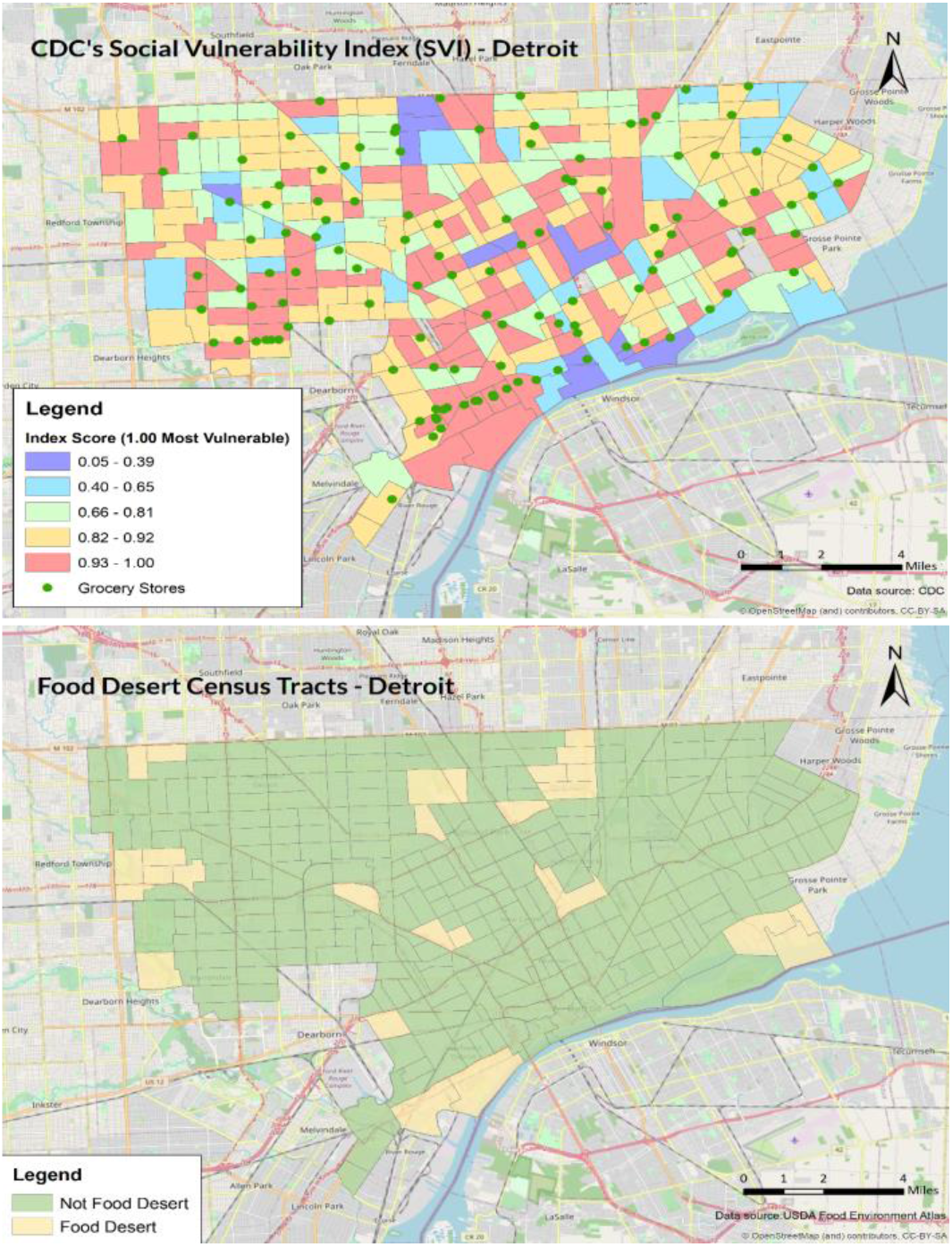
Identifying Detroit’s vulnerable population at the CT level using (*a*), 2018 CDC’s SVI figures and (*b*), 2015 USDA food desert locations

The presence of effective public transit could, however, lessen how constraining the proximity issue with food access. Transit service however was hard hit particularly during the early days of the COVID-19 pandemic. Monthly ridership analyzed over a 20-month period ranges from a low of 0.38 million in April 2020 to a high of ∼2.06 in July 2019. Figure 2 shows the magnitude of the decrease in service. The figures obtained from DDOT reveals a close to 80% decrease in April 2020 compared to the same month in the previous year. Without access to public transit, vehicle dependent households are further hindered in securing access to food.

**Figure 2:**
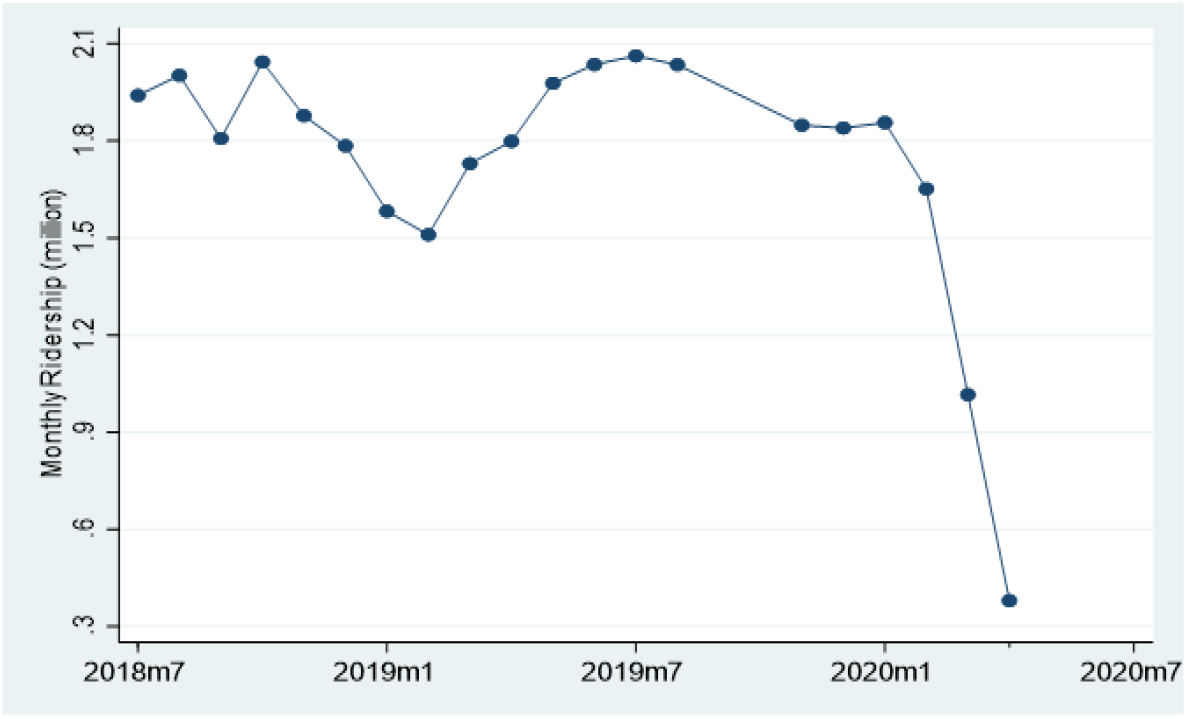
Detroit’s public transit ridership volume (in million)

In addition, we examine variables that have direct impact on food insecurity. One is looking at food insecure households based on whether the household is a SNAP recipient or not. The second acknowledges the pervasive influence of transportation on access to food by analyzing the percentage of households without a vehicle in the city. The third and the final one reflects the unique situation and the extra burden COVID-19 imposes on households given the movement from *eat in* to *order delivery* by most restaurants. We capture this by looking at the proportionate representation of households without internet access. Figures 3 and 4 provide thematic maps for SNAP recipient households and households without a vehicle at the census block group level for the city.

**Figure 3:**
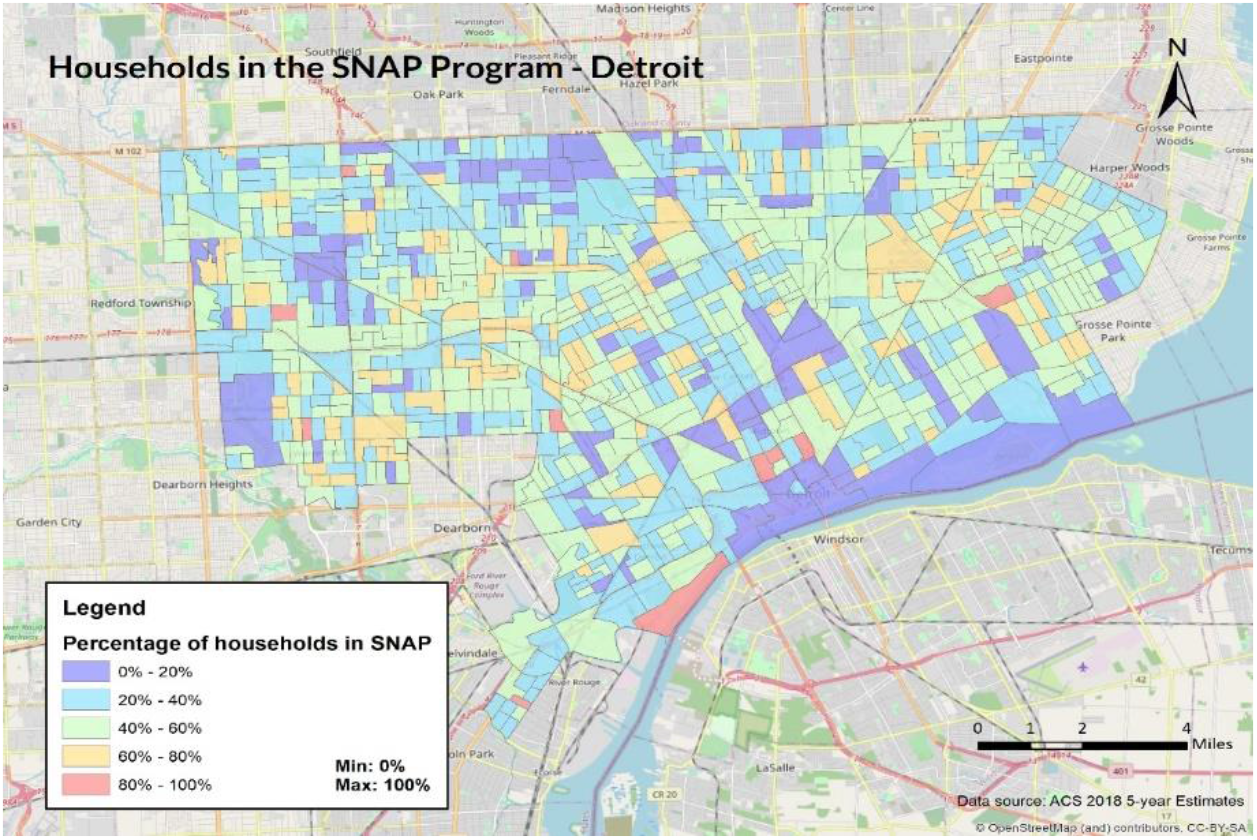
Percentage of households receiving SNAP benefits at the CBG level

**Figure 4:**
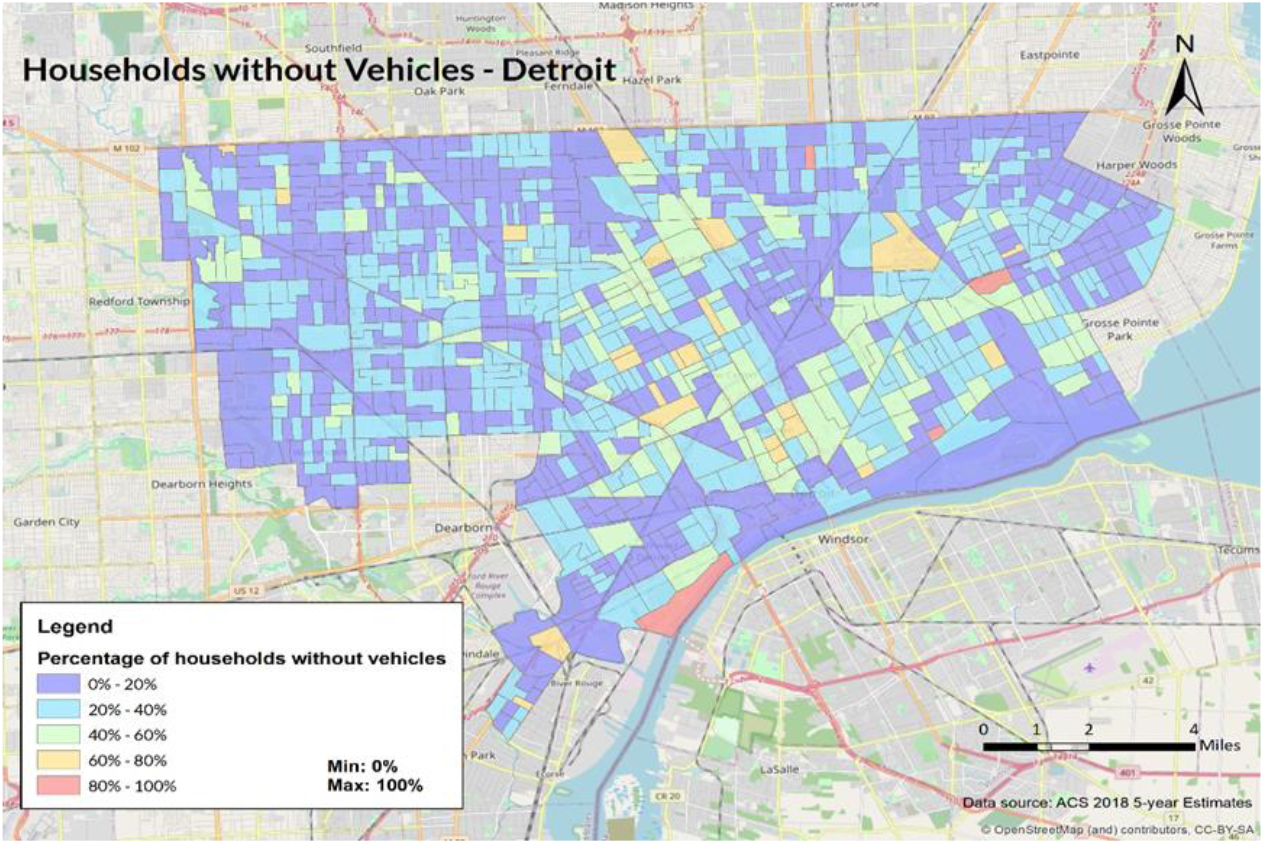
Percentage of households without vehicles at the CBG level

Figures from the 2018 American Community Survey reveal that 161,758 households, out of a total number of 676,587 households living in 1,822 census block groups in Detroit participated in SNAP. In other words, 23.9% of all Detroit households received nutrition benefits to supplement their food budget. These households spread out across the city. A block group can have as many as 940 households enrolled in the program and a percentage of participants as high as 100%. They are typically low income and unemployed households. 50% of the block groups in Detroit have a poverty rate below 10%, however, the poverty rate can be as high as 75.7%.

During the initial days of the pandemic, the city decided to partially shut down the transit system due to safety concerns, leaving 93,668 households with no vehicles in a difficult situation in terms of securing their food supply. Many families without vehicles chose to have their food delivered via a smartphone application, such as Uber Eats, Grubhub and Chowbus, or ordered their food online using a computer with internet access – an option that is only possible for households with internet access. In the City of Detroit, there are 143,654 households who do not have access to the internet. The percentage of households who have no access to the internet in a block group can vary from 0% to 100%, and the actual number of households can vary from 0 to 826.

Finally, we provide an estimate of the population of food insecure households by restricting the population to only those households that are receiving SNAP benefits, without a vehicle and without access to the internet. Using the 2014-2018 PUMS dataset, we generate a population estimate of 20,808 for all households that meet the criteria specified.

### 4.2 Assessing the severity of COVID-19 pandemic on food insecure households

The findings in this section were reported based on secondary and primary data analysis of multiple surveys conducted in Detroit during the first phase of the pandemic (March – May 2020). Although the focus of the surveys differed as did the target respondent population, they overlapped in questions regarding food and transportation security and hence provided a basis for understanding Detroit residents’ perception of food and transportation related insecurities during the pandemic as well as their overall stated travel patterns under restrictions.

Of the three surveys used in this analysis, the Detroit Metropolitan Area Communities Study (DMACS) survey was the most extensive instrument, looking into multiple behavioral aspects, including financial and health related behaviors, employment status pre and post COVID, trust in the government and media, along with questions of interest for us – insecurity related to access to food and supplies, healthcare, transportation as well as any stated changes in travel pattern. As of June 30, 2020, the survey had been carried out in three waves. Not all questions were repeated in all the waves but there was sufficient overlap for some of the questions of interest for our case study.

The second survey used in this analysis was the SNAP/EBT survey carried out by Poverty Solutions while the third survey was a short survey carried out by the Detroit Parks and Recreations Department specifically for the purpose of evaluating the food delivery pilot program. In the following subsections, we present the findings based on our secondary data analysis of the DMACS and SNAP/EBT survey while Section 4.3 presents the findings of the primary data analysis of the Detroit Food Delivery Pilot Program.

The Detroit Metropolitan Area Communities Study (DMACS) is ‘a University of Michigan initiative designed to regularly survey a broad, representative group of Detroit residents about their communities, including their expectations, perceptions, priorities, and aspirations’ [16]. DMACS had been conducting surveys of Detroit residents using a panel of respondents selected via address-based probabilistic sampling of all occupied households in the Detroit Metropolitan Area. Multiple modes were used for response collection – email, telephone interviews, mails, door-to-door campaigning etc. For the COVID-19 scenario, DMACS’ first wave of rapid response surveys was carried out between March 31, 2020 and April 14, 2020; the second wave happened between April 27, 2020 and May 7, 2020 and the third wave took place between May 28, 2020 and June 11, 2020. From information available publicly, the same respondents do not appear to be tracked throughout the three waves, although there might be overlap among respondents. The survey invitation was sent to a panel of about 1800 Detroit residents. The number of respondents in the first wave was 1044 (∼55% response rate), in the second wave was 1102 (∼61% response rate) and for the third wave was 1173 (∼66% response rate). The survey sample was weighted to represent the Detroit population.

The respondents were 77% Black, 68% younger than 55 years, 52% with a high school degree or less, and 71% with household income less than $50,000. For the first wave, we specifically looked at questions 5 and 6 of the survey that are related to concerns and behaviors during COVID-19. Question 5 states: “The COVID-19 pandemic may cause challenges for some people regardless of whether they are actually infected. How concerned are you about each of the following things?” and provided a list of options including access to healthcare, food, water and other supplies, place to live, transportation, interaction with people, getting medication and caring for family. About 82% of respondents indicated being concerned about access to food, water and other supplies while about 50% reported concerns about access to transportation. 74% indicated concerns about getting medication while 80% indicated concerns about getting healthcare as needed. By the second wave, the percentages saw a significant drop – about 53% indicated concerns with getting healthcare as needed (27 percentage point drop from first wave; p-value <0.0001) with only 39% reported concerns with getting medication (36 percentage point drop from first wave; p<0.0001). For food, water and other supplies, 77% of the respondents still reported concerns (5 percentage point drop from first wave; p <0.001) while only about 32% reported concerns with transportation access (18 percentage point drop from first wave; p<0.0001). In the third wave, 45% of the respondents reported concerns with access to food, water and supplies (32 percentage point drop from second wave) while 28% reported concerns with access to transportation (4 percentage point drop from second wave). 40% respondents expressed concerns about getting needed healthcare (13 percentage point drop from second wave) while 22% reported concerns about getting medication (17 percentage point drop from second wave). Table 1 shows the differences between the responses in different waves and their statistical significance.

**Table 1.** Summary of Respondents Concerns on Items of Interest

| Concern | First Wave<br>(Mar-Apr) | Second Wave<br>(Apr-May) | Third Wave<br>(May-Jun) |
| --- | --- | --- | --- |
| Food, water and supplies | 82% | 77%<br>(-5) | 45%<br>(-32) |
| Transportation | 50% | 32%<br>(-18) | 28%<br>(-4) |
| Healthcare | 80% | 53%<br>(-27) | 40%<br>(-13) |
| Medication | 74% | 39%<br>(-36) | 22%<br>(-17) |

*Note: Numbers in italics and in parenthesis indicate changes in percentage point in responses from the preceding survey wave*

We next looked into the self-reported travel patterns and changes as a result of COVID-19. For this, we parsed Question 6 of the survey waves 2 and 3 and a similar question that was presented as Question 3 in survey wave 3. While the underlying purpose of the question appeared to be understanding some of the safety behaviors related to COVID-19, the questions and the items were unfortunately differently worded and selectively presented in different waves, making it difficult to compare responses across the different survey waves. However, an analysis of each wave independently revealed a matched response of cancelled and not cancelled travel for any other purpose than work in the first wave – 73% respondents indicated not canceling work trips at that time. In the second wave, a single item of ‘cancelled or postponed travel’ was presented without the granularity of travel purpose and 61% responded ‘yes’ to the item. In the second wave, a new item of ‘gone to grocery shop or pharmacy’ was added which saw an 84% of respondents saying ‘yes’ to the item. The third wave did not have any of these items in the related question. One overlapping item of interest in the second and third wave was ‘Gone outside to walk, hike or exercise’ to which 62% responded ‘yes’ in the second wave and 73% responded ‘yes’ in the third wave, indicating an increase in outdoor recreational activities over time.

The third wave of survey also included questions on food availability and quality. A 10-percentage point drop could be seen in respondents agreeing that they had enough food, and the same kind of food before and after March (67% vs 57%). A 10-percentage point increase was seen in respondents indicating having enough food but not always of the same kind as before (22% vs 32%) and a 1-percentage point increase in respondents indicating not having enough to eat (9% vs 10%). For the respondents indicating not having enough food or the same kind of food, a follow-on binary response question was asked to understand the reason. 63% said ‘yes’ to stores not having stock, 46% indicated ‘yes’ to not being able to afford more food, 50% indicated ‘yes’ to being afraid to go out to buy food. However only 22% said ‘yes’ to ‘couldn’t get groceries or food delivered’ and 35% indicated having issues preventing them from going out to get food. Of the 20% respondents who indicated they received free food in the previous 7 days before taking the survey, the majority received food from food banks or pantries followed by family, friends, and neighbors and then from programs aimed at school children. Home delivered meal service like Meals on Wheels supported 14% of the respondents receiving free food. Census.gov did three rounds of national surveys with some questions on food security three of which matches perfectly with the questions asked in the third wave of DMACS survey.

Census.gov conducted 12 weeks of national surveys with questions on food security - question 25 - 27 of the questionnaire matches with the questions in the previously discussed DMACS survey, wave 3. Based on the data collected, census.gov calculated a food scarcity map for the states and for MSA areas within the states. The maps, accessible from https://www.census.gov/data-tools, shows food scarcity in the Detroit-Warren-Dearborn MSA area at the start of the survey (April 23-June 2) and at the last round of the survey (July 16-July 21). We analyzed the survey data for Detroit-Warren-Dearborn MSA for the relevant three food security related questions for the first and the last wave of the survey to understand how closely the results match our secondary data analysis of the DMACS survey as well as to understand how the food security landscape changed over the pandemic.

During the first survey, 1138 responses were recorded for Detroit-Warren-Dearborn MSA which increased to 1305 responses during the final wave. 59.4% reported having sufficient food during the first week while 63.4% reported having sufficient food during the final week of the survey. Of the responses that noted food insufficiency, 81.1% stated having sufficient food but not of the kind they like, 15.4% stated sometimes not having enough to eat and 3.5% stated often not having enough food. The corresponding percentages for the last week were 80.6%, 16.6% and 2.8%. Reasons for not having sufficient food by variety or quantity for week 1 were predominantly ‘not being able to afford’ (29%) and ‘being afraid to go out to buy’ (30%), followed by ‘not being able to get delivery’ (15%) and ‘couldn’t get out to buy food’ (10%). For the final week, the highest percentage of food insufficient respondents reported ‘not being able to afford’ as the reason (39%) followed by ‘being afraid to go out’ as the cause (33%). About 13% reported ‘couldn’t go out to buy food’ as the reason while about 6% indicated that they could not get food/grocery delivered. The percentage of respondents indicating not being able to afford sufficient food is lower in the PULSE survey compared to the DMACS survey though, from both surveys, 25-30% of residents suffered from food insufficiency because of either not being able to get food delivered or having some issues that prevented them from accessing grocery or food supply systems. In the following section we describe a food delivery program designed and implemented by the City of Detroit for its residents and assess how it met the needs of the target population.

### 4.3 Preliminary findings from the pilot program

Here we discuss the initial findings of the City of Detroit Food Delivery Pilot Program. This information is derived from two sources of information and informed by the first-hand experience of program managers. The first is the program data dashboard (N=350), using data updated as of 7/31/2020. The second is a survey (N=42) administered through the phone, two months into the program by program staff. This survey captured the experience of participants. Selection for residents surveyed was done by putting a list of participants from the program through a random number generator.

This program, since the onset of the pandemic, has allowed the City to aid 350 households; over 1,000 unique individuals, and has served over 90,000 meals. This had all been made possible through the engagement of 575 volunteers, volunteering over 6,850 delivery miles. Despite this success with limited resources, we can infer that the need within the City of Detroit is much greater. It is important to note that the City is working to scale up and serve 1,000 residents a week. Furthermore, other programs across the city, in addition to two smaller City pilots, Gleaners Direct Delivery and Brilliant Detroit Family Delivery, work to meet this need; the two smaller City pilots serve an additional 100 households a week.

From the Food Delivery Pilot Program survey, we found the following response data to be the most insightful: 86% of recipients said they have enough or more than enough food each week; 70% of food consumption by program recipients came from the delivery program (on average per week, of all food consumed by household); 69% of said they had to supplement with food from another source; 35% of recipients stated this was the first time they used a food program. From this information we can infer that, first, there are novel cases of food insecurity, given that a third of the residents stated this is the first time they used a food program. Additionally, from open-ended responses to our survey, respondents indicated that there were limitations to the food deliveries, with a lack of dairy, meat, and other refrigerated items being the largest source of these issues.

## 5 DISCUSSION OF FINDINGS

Analysis of the survey data, both secondary and primary, indicates that at the early stages of the pandemic people were apprehensive about their access to food, healthcare and transportation and a significant portion of respondents reported not having sufficient food. However, survey respondents at a later time in June-July, appear to be more confident about food sufficiency and about their access to food. Our hypothesis is that while cities and regions were not fully prepared for the intensity of the pandemic and related regulatory measures, the City of Detroit was adaptable in developing strategies and implementing them within a short period of time.

However, even with these improvements, sustainability of the pilot program is still in question. For example, facilities such as The Isaiah Food Project that are funded by private donations and staffed by volunteers, are rolling back their services. The 80% percent of respondents in the first wave who expressed concerns about access to food corroborates findings from the pilot food delivery program given that one out of every three respondents said they were using such a program for the first time. With regards to challenges with resources, it is relevant mentioning that the City’s goal of providing food delivery services to one thousand individuals each week represents only a fraction of the estimated ∼20,808 households that might benefit from the service. One way is to prioritize who gets the service based on COVID-19 status – a criteria used in the early periods of the pandemic.

The City in collaboration with other organizations developed the food delivery system which catered to the needs of the COVID positive patients and beyond. The City also deployed alternative personal transportation modes like e-scooters to provide people safe access to their destinations. In addition, since most people indicated being afraid to go out to buy food and grocery, it might be helpful to publicize the measures that have been put in place to ensure safe interactions and also to educate people about safe behaviors. A not so negligible section of people noted not being able to get food delivered as a reason for food insufficiency which may likely be due to their unfamiliarity with app based doorstep food delivery systems or not having smartphones to access those apps or being in localities not served by these services. In the future, it may be useful to develop a system to identify such localities and groups of people and collaborate with app-based delivery services to set up less technologically involved systems that can remove the access barrier.

One of the concerns from the findings is that people reported to be mobile and travel despite being COVID positive and being provided with food delivery. Reasons cited were need for baby food and special needs food as well as lack of variety in the packaged food supplied. This implies that while food delivery systems can provide a strong support to the food insecure people, it may not be completely effective in curbing trips to grocery and other food supplies. The city should plan for safe interactions at destinations like those to prevent spread of the pandemic.

## 6 CONCLUSION

The Detroit Food Delivery Pilot program was put in place to address an immediate need created by the COVID-19 crisis. Through this program, the City had aided 350 households; over 1,000 unique individuals and has served over 90,000 meals. This had all been made possible through the engagement of 575 volunteers, volunteering over 6,850 delivery miles. The rationale for our involvement in the program is to be able to provide the City with the capacity to respond to these developments in real time while creating a structure on how the responses are coordinated. We detail how the vulnerable population was identified, provide an estimate of the population and examine the severity of the pandemic on this segment of the population. The results were used to provide insights on the programmatic intervention to rectify the impact of the crisis on these individuals.

These tasks were carried out using a myriad of datasets including primary data administered on the Detroit Food Delivery Pilot program and secondary data. The program was originally designed to provide two to three weeks of contactless food delivery to households in the City of Detroit with least one individual who tested COVID-19 positive. As active infections waned and COVID-19 positive households dropped below the capacity of the program, agencies who serve other vulnerable populations were encouraged to refer families living at the intersection of food and transportation insecurity to the program.

An objective of the food pilot program is to document not only the outcome but also the process involved in modifying existing programs to better respond to a need created by the COVID-19 crisis. This allows for learnings and local adaptations by the City of Detroit as well as other cities that may be grappling with the same problem. As an example, incurring fixed costs from new acquisitions was not an option entertained due to cost issues. Instead, we explored the repurposing of existing assets, such as school buses to meet the needs. We believe that other cities could benefit from our documentation and be able to modify their program designs to better address the local peculiarities of their environment.

## Data Availability

All data used for the analysis are publicly available.

https://www.census.gov/programs-surveys/household-pulse-survey/data.html

https://www.census.gov/programs-surveys/acs/microdata.html

https://www.atsdr.cdc.gov/placeandhealth/svi/data_documentation_download.html

## ACKNOWLEDGEMENTS

This material is based upon work supported by the National Science Foundation RAPID grant under Grant No. OIA-2029518: Improving Transportation Equity to Enhance Food Security for Families Vulnerable to COVID-19. The authors are grateful to all the project team members.

## AUTHOR CONTRIBUTIONS

The authors confirm contribution to the paper as follows: study conception and design: all; data collection: all; analysis and interpretation of results: all; draft manuscript preparation: all. All authors reviewed the results and approved the final version of the manuscript.

